# Humoral and cellular immune responses to SARS CoV-2 vaccination in Persons with Multiple Sclerosis and NMOSD patients receiving immunomodulatory treatments

**DOI:** 10.1101/2021.12.22.21268127

**Authors:** H. Bock, T. Juretzek, R. Handreka, J. Ruhnau, M. Löbel, K. Reuner, H. Peltroche, A. Dressel

## Abstract

**Background:** Vaccination against SARS CoV-2 results in excellent personal protection against a severe course of COVID19. In persons with Multiple Sclerosis (PwMS) vaccination efficacy may be reduced by immunomodulatory medications.

**Objective:** To assess the vaccination induced cellular and humoral immune response in PwMS receiving disease modifiying therapies.

**Methods:** In a monocentric observational study on PwMS and patients with Neuromyelitis optica we quantified the cellular and humoral immune responses to SARS CoV-2.

**Results:** PwMS receiving Glatirameracetate, Interferon-ß, Dimethylfumarate, Cladribine or Natalalizumab had intact humoral and cellular immune responses following vaccination against SARS CoV-2. B-cell depleting therapies reduced B-cell responses but did not affect T cell responses. S1P inhibitors strongly reduced humoral and cellular immune responses.

There was a good agreement between the Interferon gamma release assay and the T-SPOT assay used to measure viral antigen induced T-cell responses.

**Conclusion:** This study demonstrates that S1P inhibitors impair the cellular and humoral immune response in SARS CoV-2 vaccination, whereas patients receiving B-cell depleting therapies mount an intact cellular immune response. These data can support clinicians in counselling their PwMS and NMOSD patients during the COVID 19 pandemic.

## Introduction

In the ongoing Covid-19 pandemic vaccination is considered the most effective measure to provide both: a significant public health benefit by slowing the spread of the virus, as well as conferring an excellent personal protection against severe COVID-19.(1, 2) While there is no evidence to suggest that SARS CoV-2 vaccination exacerbates MS activity or accelerates disability progression (3) it is known that respiratory infections are a risk factor for MS relapses. Emerging evidence indicates that COVID-19, too, has a detrimental effect on the clinical course of MS. (4, 5)

In Persons with Multiple Sclerosis (PwMS) Risk factors for severe COVID-19 have been shown to be the same as have been identified in the general population (6) Although the majority of disease modifying medications used to treat MS patients have no detrimental effect on the clinical course of COVID-19, (7-9) for B-cell depleting therapies conflicting evidence has been reported.(10, 11) It remains unresolved whether PwMS treated with these medications are at a greater risk to acquire SARS CoV-2 infection and to develop a more severe disease course. (8)

Thus, SARS CoV-2 vaccination of PwMS is broadly recommended. It remains a concern, however, whether vaccination exerts its full efficacy in PwMS receiving immunomodulatory or immunosuppressive treatment. A recent study in Israel was the first to demonstrate a reduced humoral response to vaccination against SARS CoV-2 in MS patients treated with B-cell depleting antibodies or S1P inhibitors (12), a finding subsequently confirmed by other groups. (13, 14)

However, information on the cellular immune response to vaccination in PwMS and patients with Neuromyelitis optica spectrum disorder (NMOSD is scarce. Gadanie et al found a robust cellular response in ocrelizumab treated patients.(15) Here we use routine clinical data to compare the humoral and cellular immune response to SARS CoV-2 vaccination in PwMS and NMOSD patients receiving immunomodulatory therapies to gain a better understanding of the impact of the therapy on the vaccination induced immune response.

## Methods

### Patients

In our tertiary MS center clinics we routinely determine the antibody response and the T cell response in all PwMS or Neuromyelitis optica who obtained their complete vaccination against SARS CoV-2, to allow better counselling of our patients. The use of de-identified routine clinical data from patients treated in the MS clinic of the Dept. of Neurology of the Carl Thiem Hospital in Cottbus was approved by the local ethics committee (Votum 2021-2124-BO-ff). Data of patients were included in this study if they had a negative history for Covid-19 infection and their vaccination was completed between two weeks and three months prior to blood sampling. In addition, an unaltered treatment regimen of disease modifiying therapy was required between vaccination and blood sampling. Each patient was only included once using the first data set matching the inclusion criteria. Data was obtained from 103 consecutive patients between 1st of June and the 30th of Sept. 2021. 82 patients were included in the final analysis to asses post vaccination immune responses. The study cohort consisted of 78 PwMS and 4 patients with NMOSD. Median age was 42 (range 22-70) there were 54 female and 28 male participants. The reasons to exclude patient samples is given in Fig. 1.

**Figure 1.**
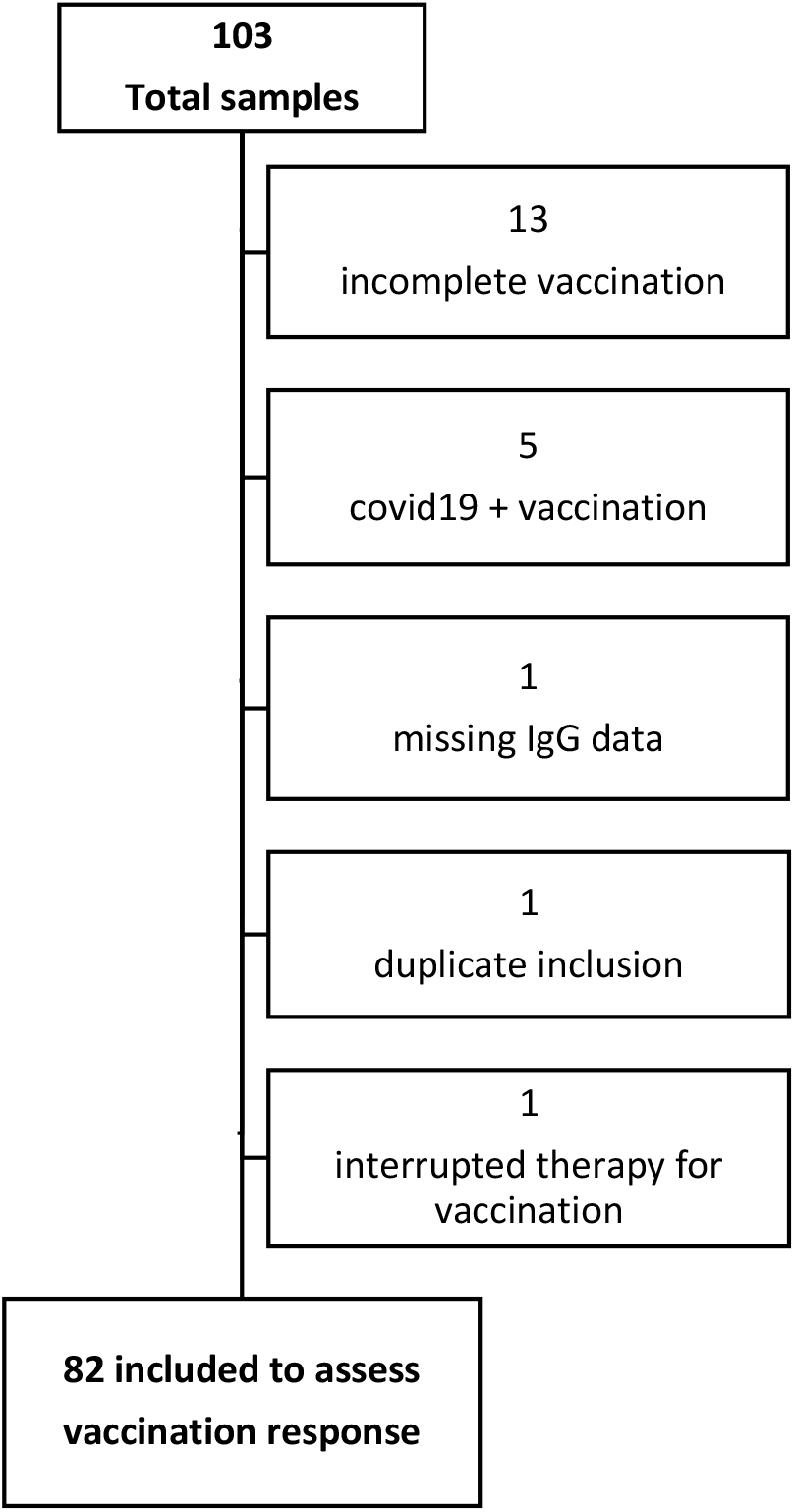
The figure shows patient inclusion and exclusion into the study.

For the comparison of the two T cell stimulations assays IGRA and T-SPOT.*COVID* all patients with data sets on both methods were included (n=91).

### Neutralizing IgG antibodies

Anti-SARS-CoV-2 IgG antibodies were determined using the LIAISON® SARS-CoV-2 TrimericS IgG assay from Diasorin. Serum or plasma was processed fully automatically on the LIAISON XL. The trimeric spike glycoprotein is the stabilized native form of the SARS-CoV-2 spike protein and a stabilized trimer can accurately detect the presence of neutralizing antibodies IgG. The cut off value is 33.8 binding antibody units/ml (BAU/ml). A higher value was determined to be positive. The clinical sensitivity is as high as 98.7% and the clinical specificity is as high as 99.5%. The test correlates with the microneutralization test: very good postive percent agreement (PPA): 100%, negative percent agreement (NPA): 96.9%. A value greater than 33.8 BAU/ml is positive. Conversion factor to neutralization assay is BAU/ml=mIU/ml times 2.6). (16) Values greater than >2080 could only be titrated late in the study or were no longer available.

### Interferon gamma response assay (IGRA)

SARS-CoV-2-IGRA from EUROIMMUN, Germany was used together with the corresponding stimulation tube set from EUROIMMUN consisting the three stimulation tubes CoV-2 IGRA BLANK, CoV-2 IGRA TUBE, CoV-2 IGRA STIM) for using with one sample. Fresh human whole blood from lithium heparin blood collection tubes is treated in the individual stimulation tubes and plasma is obtained from this. The protocol followed the manufacturer’s instructions. Plasma not immediately processed was stored cell-free under −17° C for a maximum of 2 weeks. The concentration of released interferon-gamma in the plasma is then determined. The interferon-gamma concentration in the plasma of the BLANK represents the individual interferon-gamma background and was subtracted from the interferon-gamma concentration of the plasma, in tubes TUBE and STIM. After BLANK subtraction, the interferon-gamma concentration in the STIM condition must still be higher than the BLANK value itself in order to ensure a sufficient number and stimulability of the immune cells. Cob’centrations greater than 2500IU/µl were not titrated in the clinical routine analysis. The manufacturer defines values greater than 120 IU/µl to be stimulable by SARS-CoV-2 antigens values below 100IU/µl are considered negative, values in between are borderline results.

### T-SPOT.*COVID*

T-cell mediated immune response to SARS-CoV-2 vaccination (and/or infection) was determined using the T-SPOT.*COVID* (Oxford Immunotec). The Enzyme Linked ImmunoSpot enumerates the CD4 as well as CD8 T-cells that respond to stimulation with antigens of SARS-CoV-2 by secretion of interferon-g (INF-g). This is immobilized on the bottom of the microtiter by INF-g-specific antibodies. In the development step each spot corresponds to one activated T-cell. The assay was performed according to the manufacturers’ instruction. 2.5×10^5 peripheral blood mononuclear cells (PBMC) were seeded into each of four microtiter wells for the nil control, the spike protein (S1) stimulation, the nucleocapsid stimulation and the positive control. The test is considered positive if at least one stimulation shows 8 or more spots (more than the nil control), negative if no stimulation produced more than 4 spots and borderline for all other spot constellations.

Current vaccines induce serologic responses to the spike protein, while an infection exposes the immune system to the inner proteins as well. Thus, a positive spike- and a negative nucleocapsid-stimulation indicates vaccination, whereas an infection results in positive results to both antigens.

### Statistics

For visualization and statistical analyses Graphpad Prism 8.2 (GraphPad Software Inc.) was used. Normal distribution of data was assessed using the Shapiro Wilk test. Since not all data passed the normality test more than two groups were compared using the Kruskal-Wallis test followed by Dunn’s multiple comparison test. For all analyses a p-value < 0.05 was regarded as significant.

## Results

### Comparison of the IGRA and T-SPOT.*COVID* assays

Both assays rely on the viral antigen induced induction of IFN-g secretion in T-cells. While the T-SPOT assay is a semiquantitative assay that is based on the counting of individual T-cells that respond to antigenic stimulation, the IGRA quantifies the total amount of IFN-g released from all cells in the sample upon challenge with viral antigens. In our cohort there was a 79% agreement (70 of 91 sample pairs) between both methods. Converting results considered “borderline” according to the manufacturers instruction into “positive” increased the agreement between both methods to 88% (80 of 91 sample pairs). (Table 1)

**Table 1.** Comparison of IGRA and T-SPOT.*COVID* Results to Determine Cellular Immune Responses

|  |  | T SPOT.COVID |  |  |  |
| --- | --- | --- | --- | --- | --- |
|  |  | neg | pos | border* | total |
| IGRA | neg | 10 | 3 | 0 | 13 |
|  | pos | 6 | 62 | 5 | 73 |
|  | border* | 2 | 3 | 0 | 5 |
|  | total | 18 | 68 | 5 | 91 |

\* borderline results as defined by the manufacturer

### IgG responses

Vaccination induced spike-protein-specific IgG responses in patients treated with Interferon-ß (IFN-ß), Glatirameracetat (GLAT), Dimethylfumarate (DMF), Natalizumab (NAT), Cladribine (CLAD), Alemtuzumab (ATZ) were indistinguishable from IgG responses elicted in untreated MS patients. In agreement with previous studies the IgG responses observed in patients receiving S1P inhibitors or B-cell depleting therapies were strongly diminished. (Fig. 2a) While the IgG titer reflecting protective immunity against Covid 19 remains unknown, the assay cut off indicating a spike-protein-specific IgG response is 33.8 BAU/ml. Using this cut off, B-cell responses were detectable in 9 of 16 ocrelizumab 2 of 4 Rituximab and 1 of 1 ofatumumab treated patient. (Fig. 2 a and Tab. S1). All PwMS who had detectable CD19+ B-cells in the peripheral blood in FACS analysis mounted a vaccination induced IgG response whereas there was a great heterogeneitiy of the IgG response in PwMS with complete depletion of B-cells from the peripheral blood. (Tab S1).

**Figure 2.**
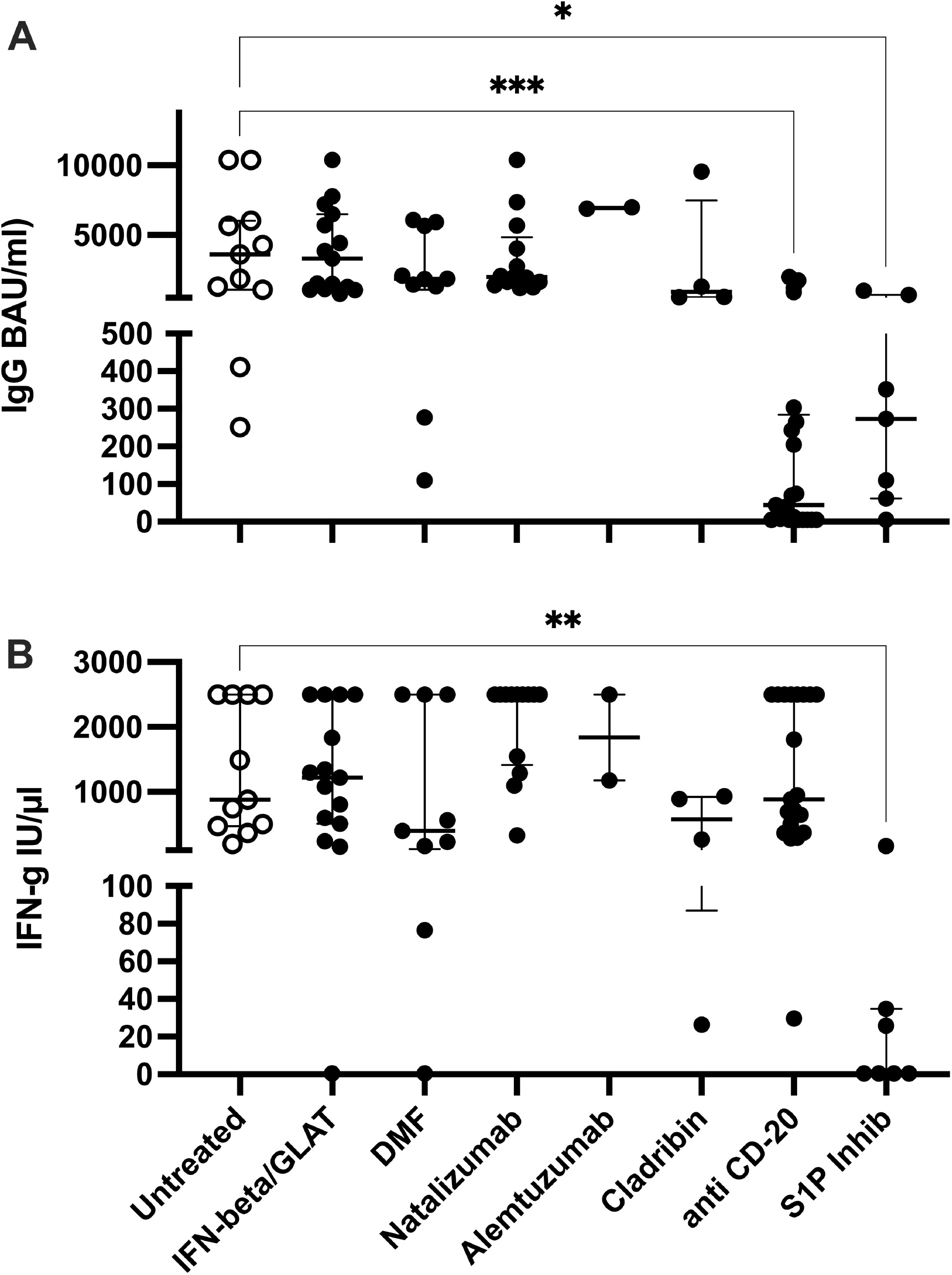
Spike protein specific IgG production (A) and SARS CoV-2 specific IFN-g production (B) in PwMS and NMOSD patients. Horizontal bars indicate median. A p<0.05 was considered statistically significant. * p<0.05; ** p< 0.01; ***p< 0.001

Under S1P treatment B-cell responses were detectable in 4 of 5 fingolimod and 1 of 1 ozanimod and 1of 1 siponimod treated patients. Although the absolute number of patients in this subgroup is small, in fingolimod treated PwMS higher lymphocyte counts were associated with IgG titers. (Tab. S1)

### T –cell responses

IFN-ß, GLAT, DMF NAT or ATZ or CLAD did not affect the cellular immune response to vaccination as measured using the IGRA. The two patients with negative IGRA results were clearly positive in the T-spot test (data not shown). Furthermore, B-cell depletion in PwMS which inhibited the humoral response had little or no effect on the cellular immune response. Upon stimulation IFN-g release was detectable in all but one patient, who, interestingly, had a weak but detectable IgG response. (Fig 2b and Tab. S1) In contrast, S1P inhibition resulted in an almost complete absence of T-cell responses in both IGRA and T-SPOT assays. The only PwMS treated with S1P inhibitors who developed a weak T-cell response had been vaccinated with Johnson and Johnson. (Tab. S1)

## Discussion

We utilized routine clinical data to investigate the humoral and cellular responses of PwMS and patients with NMOSD to SARS-CoV-2 vaccination in a single center observational study. To determine cellular responses, we used two independent methods. The IGRA determines the overall IFN-g production in stimulated blood samples and provides easily quantifiable data, whereas in the T-SPOT assay the number of T-cells responding to a specific antigen is counted. Since, the T-SPOT assay uses the spike protein and the capsid antigen in two separate stimulations, it can be used to distinguish between immune responses induced by Covid-19 infection and those elicited by vaccinations. Our data demonstrates good agreement between both assays, suggesting that either one is suitable to determine T-cell responses in clinical routine diagnostics.

Our results indicate that the majority of immunomodulators used to treat PwMS have no effect on the immune response to SARS-CoV-2 vaccination and on average patients treated with these medications developed the same humoral and cellular response levels as the untreated control group. In contrast, we detected significantly impaired humoral responses in patients treated with B-cell depleting therapies or with S1P inhibitors. Unfortunately, the latter group also showed a lowered or absent T-cell response. This observation is in good agreement with previous reports (12-14).

Importantly, despite being weaker than in untreated PwMS, IgG responses in 6 out of 7 patients treated with S1P inhibitors and 10 out of 17 patients treated with ocrelizumab remained above the assay cut-off, indicating a weak but detectable humoral immune response. While it is not known which IgG levels reflect protection against SARS-CoV-2 infection or severe disease courses, it is likely that the very low titers observed in these patient groups are not protective.(17) Consequently, the ability of these patients to mount an immune response suggests that they may benefit from additional booster vaccinations. This hypothesis is supported by a study investigating the B cell response to booster vaccination in Rituximab treated rheumatologic patients (18).

In contrast to the impaired humoral response our results demonstrate an intact T-cell response to SARS-CoV-2 vaccination in PwMS and NMOSD patients treated with B-cell depleting antibodies. This observation is in agreement with two very recent studies investigating T-cell responses in vaccinated MS patients. (19, 20)

Although the number of participants in this observational monocentric study is relatively small for each treatment group, our key results are statistically significant. Another limitation of this investigation is that no data on the pre-vaccination immune-status is available and thus, previous exposure to SARS-CoV-2 may have occurred. However, apart from PwMS who were excluded for their known history of COVID disease, no other study participant showed an anti-capsid T-cell response in the T-SPOT assay, which exclusively indicates antigen contact by infection. Since, the vast majority of our patients received mRNA based vaccines, it would be of great interest to obtain similar data on PwMS who received vector based vaccines.

The data presented in this study provides clear evidence on the cellular and humoral immune responses in MS and NMOSD patients receiving disease modifying therapies. With the notable exception of S1P inhibitors no treatment investigated inhibited the cellular immune response. Furthermore, the majority of patients treated with S1P inhibitors and half of the patients receiving B cell depleting therapies did show a low but detectable vaccination induced SARS-CoV-2 spike protein specific IgG production. This leaves only a small group of patients receiving S1P inhibitors for whom we could detect no immune response after vaccination.

We believe that these results may aid clinicians in their decision to select the best immunomodulatory treatments for PwMS under the circumstances of the pandemic and to make informed decisions on the potential benefit of additional vaccinations.

## Data Availability

All data produced in the present study are available in deidentified form upon reasonable request to the authors

