## Supplemental Table 1 for "Humoral and cellular immune responses to SARS CoV-2 vaccination in Persons with Multiple Sclerosis and NMOSD patients receiving immunomodulatory treatments"

| PAT_SEX | PAT_AGE | Treatment | Days last infusion to vaccination | SARS2-IgG_TITER_NUM | IGRA_TEST IFN-g (IU/ml) | T-SPOT_RESULT | Vaccination | Lymphocytes. (GPT) | CD19+B-cells (cells/µl) |
|---|---|---|---|---|---|---|---|---|---|
| B-cell depleting treatment | | | | | | | | | |
| m | 45-49 | Rituximab | 381,00 | 5 | 2500 | POSITIV | Biontech | 0,96 | 0 |
| f | 50-54 | Rituximab | 99,00 | 5 | 367 | POSITIV | Johnson&Johnson | 1,61 | 0 |
| f | 55-59 | Rituximab | 132,00 | 74 | 2500 | POSITIV | Biontech | 1,05 | 0 |
| m | 40-44 | Rituximab | 126,00 | 243 | 2500 | NEGATIV | Biontech | 1,38 | 0 |
| f | 35-39 | Ocrelizumab | 149,00 | 5 | 503 | POSITIV | Biontech | 2,19 | 0 |
| m | 40-44 | Ocrelizumab | 553,00 | 5 | 948 | POSITIV | Biontech | 1,15 | 0 |
| f | 35-39 | Ocrelizumab | 127,00 | 5 | 2500 | POSITIV | Biontech | 2,79 | 0 |
| f | 40-44 | Ocrelizumab | 331,00 | 5 | 1809 | POSITIV | Biontech | 1,25 | 0 |
| f | 35-39 | Ocrelizumab | 199,00 | 5 | 721 | POSITIV | Biontech | 1,79 | 0 |
| f | 40-44 | Ocrelizumab | 120,00 | 8 | 648 | POSITIV | Biontech | 1,6 | 0 |
| f | 35-39 | Ocrelizumab | 127,00 | 12 | 2500 | POSITIV | Moderna | 1,69 | 0 |
| m | 40-44 | Ocrelizumab | 164,00 | 39 | 2500 | POSITIV | Biontech | 2,69 | 0 |
| f | 30-34 | Ocrelizumab | 158,00 | 70 | 30 | NEGATIV | Biontech | 0,98 | 0 |
| m | 20-24 | Ocrelizumab | 184,00 | 205 | 2500 | POSITIV | Biontech | 1,18 | 0 |
| m | 40-44 | Ocrelizumab | 178,00 | 265 | 2500 | POSITIV | Biontech | 3,26 | 0 |
| f | 40-44 | Ocrelizumab | 142,00 | 303 | 372 | intermediate | Biontech | 2,53 | 554 |
| f | 30-34 | Ocrelizumab | 134,00 | 907 | 285 | POSITIV | Biontech | 2,68 | 0 |
| m | 60-64 | Ocrelizumab | 208,00 | 1200 | 293 | POSITIV | Biontech | 1,28 | 0 |
| f | 40-44 | Ocrelizumab | 121,00 | 1740 | 888 | POSITIV | Moderna | 1,61 | 18 |
| f | 25-29 | Ocrelizumab | 159,00 | 2000 | 695 | NEGATIV | | 2,51 | 134 |
| f | 30-34 | Ofatumumab | n.a. | 44 | 335 | POSITIV | Johnson&Johnson | n.d. | 0 |
| S1P modulating Treatment | | | | | | | | | |
| m | 45-49 | Fingolimod | n.a. | 5 | 1 | NEGATIV | Biontech | 0,25 | n.d. |
| f | 40-44 | Fingolimod | n.a. | 62 | 1 | n.d. | Biontech | 0,39 | n.d. |
| f | 20-24 | Fingolimod | n.a. | 273 | 26 | n.d. | Moderna | 0,32 | n.d. |
| f | 30-34 | Fingolimod | n.a. | 709 | 35 | NEGATIV | Biontech | 0,42 | n.d. |
| f | 65-69 | Fingolimod | n.a. | 997 | 1 | NEGATIV | Biontech | 0,44 | n.d. |
| m | 60-64 | Siponimod | n.a. | 352 | 0,5 | NEGATIV | Biontech | 0,29 | n.d. |
| f | 40-44 | Ozanimod | n.a. | 110 | 165,7 | POSITIV | Johnson&Johnson | 0,52 | n.d. |
